# Continuous Multimodal AI with Wearable Vital Signs Predicts Postoperative Complications in the General Ward

**DOI:** 10.1101/2025.11.25.25340950

**Authors:** Robin P. van de Water, Axel Winter, Daniela Zuluaga Lotero, Bjarne Pfitzner, Lara Faraj, Bert Arnrich, Patrick Rockenschaub, Felix Krenzien, Thomas Malinka, Wenzel Schöning, Christian Denecke, Johann Pratschke, Igor M. Sauer, Max M. Maurer

## Abstract

Surgery is inherently associated with complications, making early detection the cornerstone of timely intervention and improved outcomes. Artificial intelligence (AI) has been shown to predict severe events such as sepsis and mortality after surgery within intensive care units (ICUs). However, most complications occur on general wards (GW), where staffing and technical monitoring constraints impede effective real-time detection of complications. Here, we present a real-time, multimodal AI-based complication prediction system for surgical GWs, combining routine clinical data with continuous high-resolution vital signs derived from telemetric photopletysmography (PPG) sensors into digital patient representations. A total of 1,285 patients undergoing esophageal, gastric, liver, pancreatic, and colorectal surgery were prospectively enrolled. Baseline patient characteristics, intraoperative data, ICU parameters, and GW data, including 270,603 hours of recorded telemetric vital signs, were used to detect postoperative intra-abdominal infections. We demonstrate a high median area under the receiver operator characteristic (AUROC) of 0.90 (0.89-0.91) for the detection of surgery-related infections. Complications could be predicted 9 hours in advance with only a minor reduction of the AUROC: 0.89 (0.88-0.89). Continuous wearable data increased the AUROC by 8% and the Area Under the Precision-Recall Curve (AUPRC) by 109%, outperforming other modalities in our ablation experiments. Further development into an AI-based alarm system outperforms traditional early warning scores. These findings highlight the potential of high-dimensional, multimodal, real-time risk stratification to support earlier detection of adverse events in surgical patients. Our results reveal continuous monitoring, using minimally intrusive vital signs, as a key component of an intelligent, data-driven smart ward.

## Introduction

More than 300 million surgical procedures are performed worldwide each year^1^, linking one-third of all hospital costs to surgical interventions^2^. According to recent findings in the Western population, one out of two individuals undergoes abdominal surgery in their lifetime^3^. Despite considerable advances in surgical techniques, more than 25% of the patients who undergo major abdominal surgery experience a life-threatening event during hospitalization^4^. Postoperative complications are therefore a pivotal determinant for surgical outcomes, leading to higher mortality, reduced disease-free survival, impaired quality of life, and increased costs for society^5–11^.

Early detection of postoperative complications is a cornerstone of failure-to-rescue strategies as it enables immediate therapeutic intervention. Yet, rapid recognition of complications remains challenging as physiological patterns are often subtle, nonspecific, and time-varying, with substantial heterogeneity between patients^12^. By consequence, patient deterioration is often detected only after a self-reinforcing cascade of aggravation has already begun.

Artificial intelligence (AI) has been shown to predict complex severe adverse events, such as cardiac arrest or sepsis, in recent pilot studies performed in high-technology intensive care units (ICUs)^13,14^. However, in the field of general surgery, most complications emerge only after the patient has been transferred to the general ward (GW). Compared to the ICU, GW care is constrained by lower medical staffing, limited monitoring equipment, and less continuous clinical surveillance. In particular, vital-sign monitoring is often confined to intermittent spot checks, resulting in long intervals of insufficient observation that prevent early detection^15^.

We hypothesized that incorporating telemetric monitoring into digitalized clinical data would yield accurate AI-driven digital patient representations, enabling early identification of clinically relevant complications after major gastrointestinal, hepatobiliary, and pancreatic surgery. We prospectively developed multimodal AI prediction models for surgical ward patients, serving as a scalable foundation for future semi-autonomous care frameworks. The System integrates routinely generated data — including patient characteristics, intraoperative parameters, ICU data, laboratory values, semantic clinical notes, and medications — with high-resolution telemetric vital signs to create a continuous digital real-time representation of the patient’s physiological state. Aiming to detect clinically relevant yet highly granular complications, these include deep surgical site infections (SSI-III)^16^, bile leakages^17^, and pancreatic fistulas^18,19^. We translated our model into an actionable alarm framework for medical staff to improve failure-to-rescue management on GWs.

## Results

A total of 1,285 patients were prospectively enrolled in the Clinical ASSist and Alert Algorithms (CASSANDRA) study at a tertiary, high-volume surgical center in Germany between 2022 and 2025. After applying exclusion criteria, we retained a cohort of 1.136 subjects for machine learning (ML) development. Table 1 describes characteristics of the selected patient cohort.

**Table 1:** *Description of the collected data.* Numeric variables are summarized by *median [IQR]*. Categorical variables are indicated as percentages of the entire cohort (%).

| Characteristic | Value | Surgery System† | Amount |
| --- | --- | --- | --- |
| **Years collected** | 2022–2025 | Upper GI | 50 (4.4%) |
| **Average data points per patient** | 673,881 [455,829, 1,021,370] | Colorectal | 336 (29.6%) |
| **Number of patients** | 1,136 | Liver | 472 (41.5%) |
| **Age at admission (years)** | 63.0 [53.0, 71.0] | Pancreas | 277 (24.4%) |
| **Female** | 455 (40.1%) |  |  |
| **SSI-III** | 111 (9.8%) | Wearable data‡ |  |
| **Of which occurred on GW** | 96 (86.5%) | Wrist monitor | 1104 (97.2%) |
| **Time to SSI-III\* (days)** | 6.9 [4.0, 8.0] | Corsano Band 1 | 331 (29.1%) |
| **Hospital death** | 8 (0.7%) | Corsano Band 2 | 773 (68.0%) |
| **Hospital length of stay (days)** | 9.9 [6.2, 18.3] | Core Device | 1047 (92.2%) |
| **ICU length of stay** | 1.4 [0.9, 2.9] |  |  |
| **GW length of stay** | 6.0 [4.1, 10.3] |  |  |
| **No. of variables** | 7,280 |  |  |

\*From the moment of admission.
†Patients can undergo surgery for multiple systems; here, the primary surgery system is referred to.
‡Due to logistic issues in data collection, not every patient has usable wearable data; see discussion. Description of the collected data. Numeric variables are summarized by median percentiles: [0.25, 0.75]. Categorical variables are summarized by incidence (%).

Digital patient representations were synthesized, spanning the entire perioperative journey from preoperative patient baseline data to discharge or the first occurrence of the designated complication. Telemetric vital sign monitoring included a wrist-based PPG sensor with an accelerometer, as well as body core temperature sensors attached to the chest. The representations integrate heterogeneous data using early multimodal fusion across multiple hospital information systems, including demographics and comorbidity profiles; procedural and intraoperative parameters; postoperative ICU vital-sign data; medication and fluid administrations; laboratory results; and free-text clinical notes. Data was transformed into static covariates and intermittently aggregated time series.

The full study dataset included 5.4 million ICU measurements, 2.5 million laboratory values, and 13.8 billion PPG data points collected continuously at 32 Hz. After preprocessing, the data were segmented into 30-minute intervals, resulting in 7.280 total extracted features per segment for each patient. Multiple model architectures were evaluated, including Logistic Regression, Random Forest, Gradient Boosted Trees: XGBoost^20^, LightGBM^21^, and CatBoost^22^; and Deep learning: Transformer^23^, Gated Recurrent Unit (GRU)^24^, and a Temporal Convolutional Network (TCN)^25^. Area Under the ROC Curve (AUROC), Area Under the Precision–Recall Curve (AUPRC), and fast training times were our evaluation criteria. LightGBM was selected as the primary model on this basis. We created two models: CASHEWS (*Clinical ASsist Heuristic Early Warning System*) and CASHEWS+, which also integrates data from wearable monitoring devices. A prediction horizon of 9 hours was chosen, as it strikes a balance between good performance and sufficient time to intervene. We evaluate our approach in comparison with an existing non-ML risk-stratification metric, the Remote Early Warning Score (REWS)^26^.

### Dynamic risk stratification for GW complications

Figure 2a (AUROC) and 2b (AUPRC) present the prediction performance of models built at successive stages along the perioperative patient trajectory. Preoperative information alone failed to predict postoperative complications, as indicated by an AUROC of around 0.50. Similarly, adding intraoperative data and subsequently ICU features did not improve predictability. By contrast, the integration of routine GW into a dynamic model (CASSHEWS) demonstrated a considerable increase. The integration of continuous telemetric vital sign monitoring (CASSHEWS+) resulted in an 8 percent increase in AUROC and a 109 percent increase in AUPRC. Applying the calculated telemetry-based REWS yielded substantially lower results (AUROC: 0.55; AUPRC: 0.02). Note that AUPRC results are driven by prevalence, which is on average 0.12 for the Pre-/Intra-/Post-operative models and 0.006 for the dynamic models.

**Figure 1:**
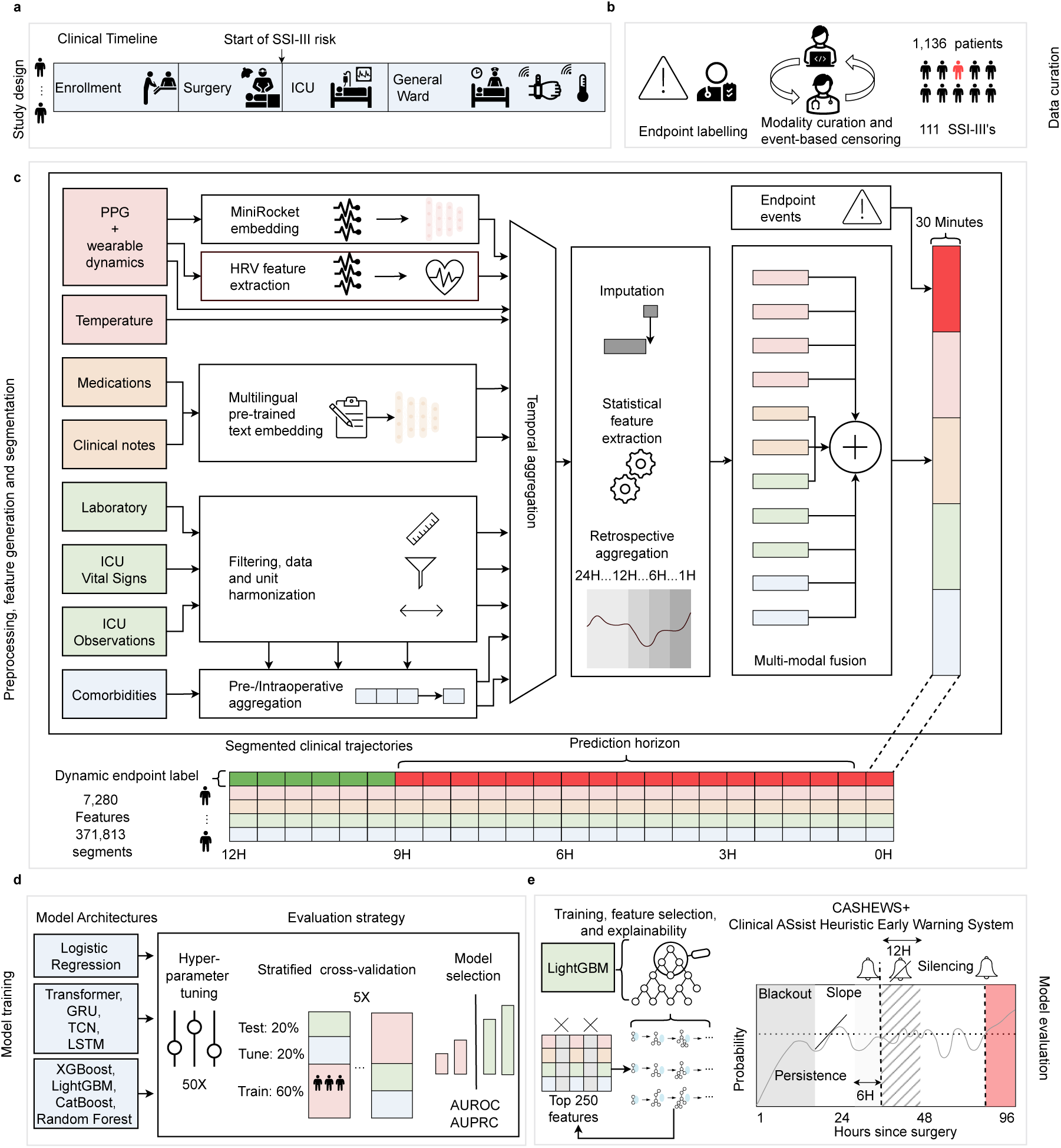
Our data segmentation and aggregation pipeline for training CASHEWS+. a: The patient journey of surgical patients. **b:** Data curation is done by precise endpoint labeling and extraction from 4 patient information systems**. c:** Segmented clinical trajectory creation: Data from the different modalities is processed in a proprietary manner. PPG data is fed into the time-efficient MINIROCKET embedding model, and Heart Rate Variability (HRV) features are extracted from time-domain and frequency-domain data. Clinical texts, medications, and fluids are embedded using a pre-trained, multi-lingual JinAI model^27^, and common Natural Language Processing (NLP) features were extracted. The clinical data is filtered, harmonized, and cleaned. Curated comorbidities are also used, and all clinical features are aggregated to create data from before the monitoring period. Then, all data within this time interval were aggregated, missing value imputation was performed, statistical features (skew, entropy, standard deviation, minimum, and maximum) were extracted, and data from previous timesteps were aggregated to ensure that data dynamics were captured appropriately. Multi-modal fusion and endpoint insertion result in segmented clinical trajectories with a prediction horizon of 9 hours. **d:** Model training: First, a range of common architectures were tested through tuning hyperparameters and performing cross-validation, after which the best performing model was chosen in terms of predictive performance and speed: LightGBM. This model architecture was then evaluated for the best features through Shapley explanations. The resulting model serves as an alarm system, featuring mechanisms to prevent erroneous evaluations. Specifically, a “blackout” prevents activation within the first 8 hours. The “slope” coefficient and “persistence” are used to quantify an increase in the probability of sounding an alarm. An 8-hour “silencing” period is implemented to prevent alarm fatigue.

**Figure 2.**
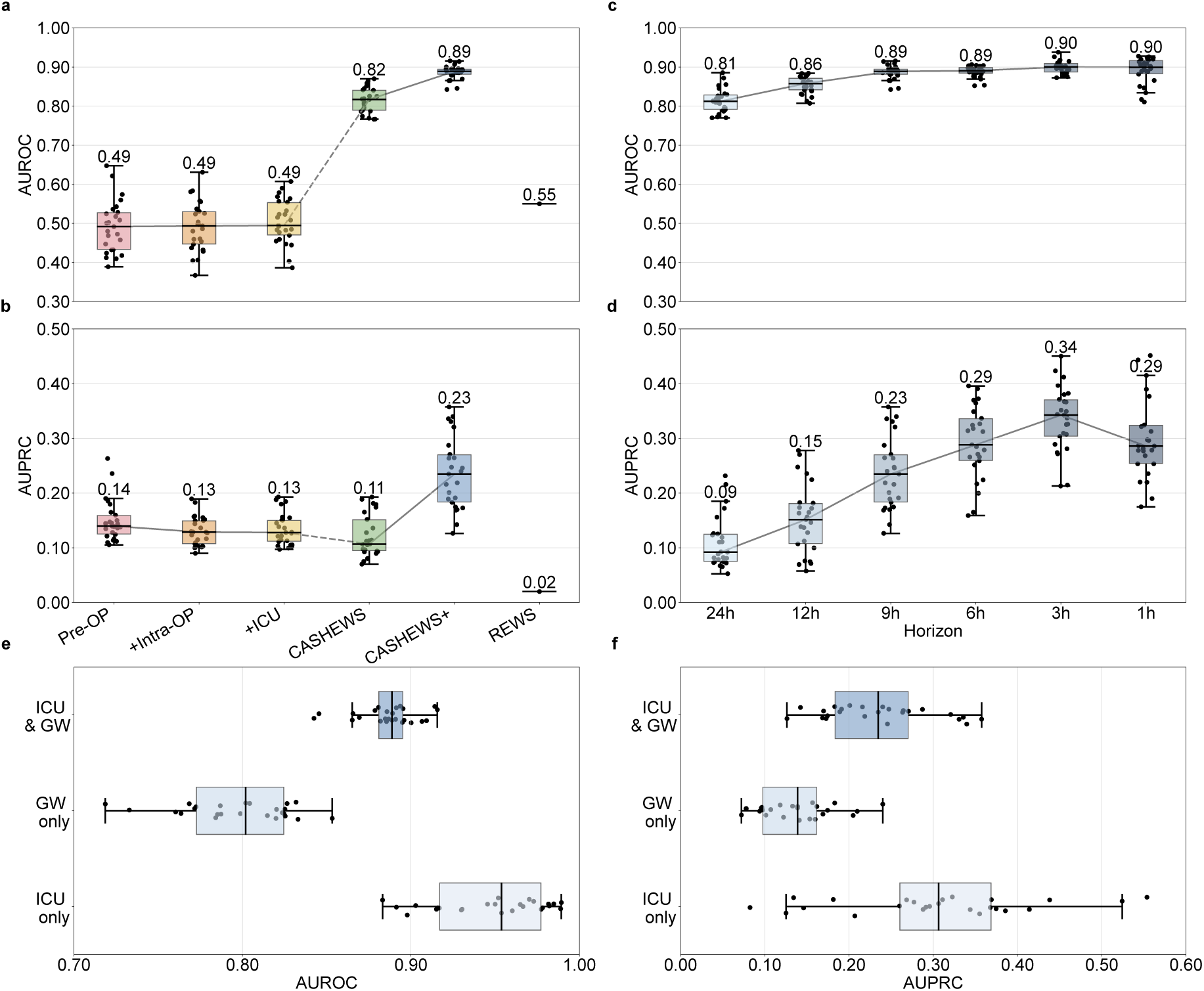
Model progression performance comparison. **a)** The Area under the receiver operating curve (AUROC) and **b)** patient-level area under the precision recall curve (AUPRC) model progression, from left to right: pre-operative static variables (Pre-OP), variables including the surgical procedure (+Intra-OP), variables including the ICU stay (+ICU). The second group of methods is dynamic (real-time) with a horizon of 9 hours: the dynamic (online prediction) model without wearable data (CASHEWS), the complete model with the 500 most important features selected (CASHEWS+), and the remote early warning score (REWS)^26^. The online prediction systems are calculated with snoozed episode-level precision-recall^28^. **c), d)** Comparison of the time horizon for CASHEWS+ for the detection of our endpoints. Here, the last hour was set to be positive, and all subsequent time windows were removed from training and evaluation. **e), f)** AUROC and AUPRC, respectively, of model experiment variations: using both GW and ICU segments, using GW time segments for testing (GW only), and only using ICU segments for testing.

Evaluating the detection at several points in time prior to the event of interest, the highest detection performance was observed between an hour and three hours preceding the clinical recognition of the complication, with a detection rate of 0.90 ± 0.03 and 0.90 ± 0.01 (median ± standard deviation), respectively. Analysis of segments up to 24 hours before detection revealed a steady increase in accuracy leading up to the event. In this context, prediction at a 24-hour lead time still achieved an AUROC of 0.81 ± 0.03.

Finally, the bottom row demonstrates that predicting endpoints solely in the ICU, despite the lower prevalence, is considerably easier for CASHEWS+. The evaluation on only the GW segments still performs reasonably (AUROC 0.81 ± 0.03). This result suggests that we might benefit from even more continuous monitoring capabilities on the general ward and that detecting the endpoint is non-trivial in a mobile ward.

### Modality ablations

The impact of each modality on prediction performance was evaluated by ablation of individual modalities without any feature selection, as shown in Figure 3. Relative to the base model with all 7,280 features, removing aggregated data, static comorbidities, and text embeddings resulted in minimal performance degradation. ICU data, temporal features, and laboratory values had a marginal impact on model performance. In contrast, removing wearable data had a major impact in terms of AUROC (9%) and AUPRC (50%). To determine the contribution of individual wearable processing strategies, we performed additional ablations, removing each signal-processing approach in all general ward patients. While the addition of each wearable modality has an impact with varying interquartile ranges, adding at least one wearable modality improves performance in every case. Moreover, the Precision-Recall and Receiver Operating Characteristic curves demonstrate the precise performance improvement of the wearable data: CASHEWS+ over CASHEWS. The REWS^26^ rule-based baseline performs especially poorly compared to our ML-driven system.

**Figure 3:**
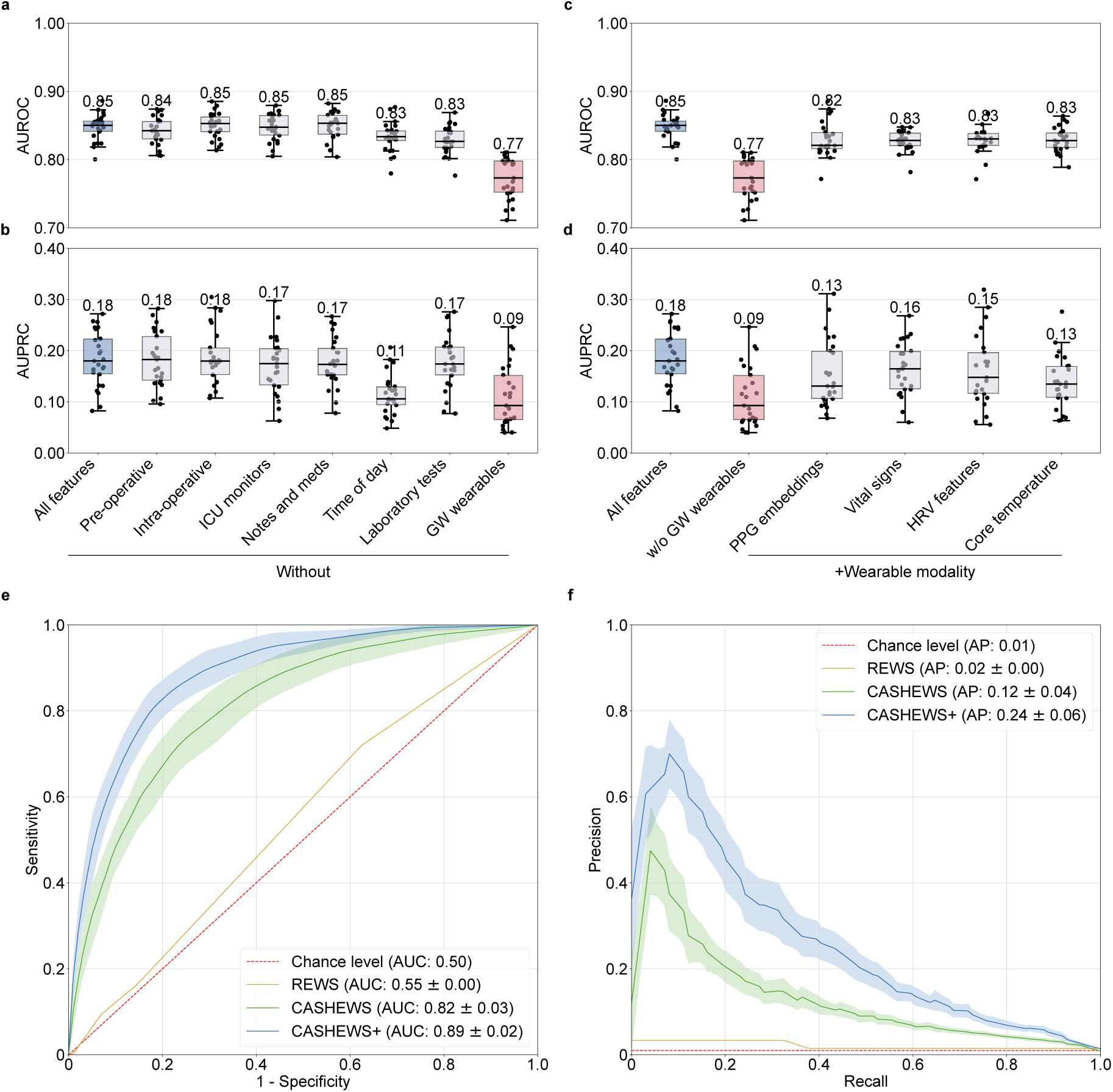
Modality ablation and prediction performance curves. **a)** AUROC and **b)** AUPRC boxplots for models after ablation of distinct modalities. Here, we refrain from feature selection to more accurately distinguish the effect of excluding features. **c)**, AUROC, and **d)** AUPRC boxplots for models after the addition of individual wearable modalities to the model without GW wearables. **e)**, Receiver Operating Curve and Precision Recall Curve comparison; CASHEWS: Clinical ASsist Heuristic Early Warning System; CASHEWS+: dynamic prediction model with wearable monitoring. Baseline: Post-ICU static endpoint prediction based on ICU data, pre-and intra-operative features

### Explanation analysis

The single feature weights of the complete model, as well as the most important modalities according to SHAPley^30^ explainability analysis, are shown in Figure 4. Body-temperature entropy emerged as the most impactful individual physiologic feature, followed by respiration entropy and mean HRV (Figure 4b). Among laboratory results, Mean Corpuscular Hemoglobin, C-reactive protein, and Lipase were the most important model components (Figure 4c). Within ICU data, the fraction of inspired oxygen ranked highest (Figure 4d). Figure 4e shows that laboratory features contribute the largest SHAP values overall, closely followed by wearables. Finally, with the t-SNE^29^ visualization of the full dataset as the mean of all feature values (Figure 4f), we demonstrate the (diss)-similarity of the modalities and individual features. The separate clusters suggest that wearable features and lab features contribute significantly to the diversity of the feature space.

**Figure 4.**
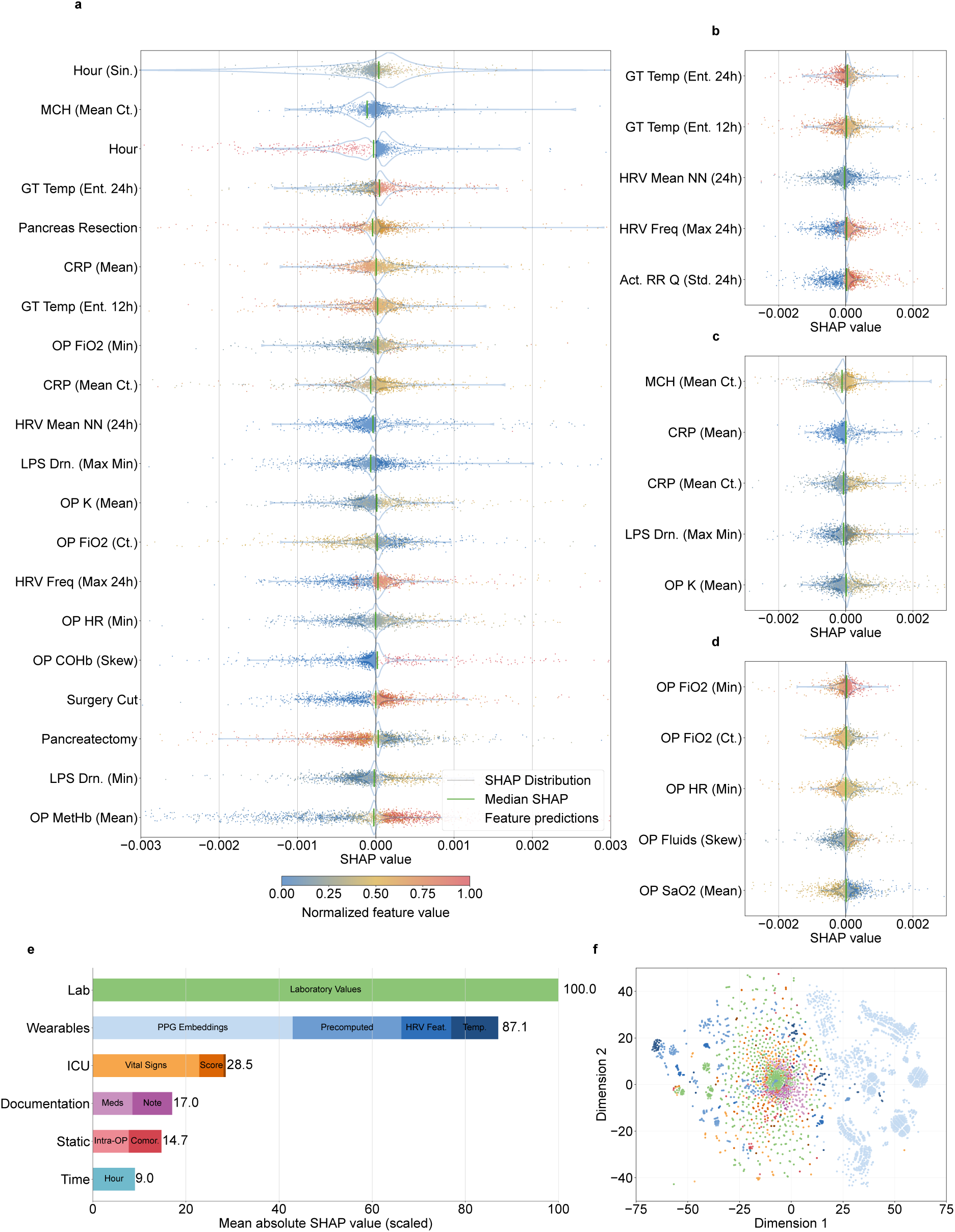
SHAP-based feature importance visualized. **a)** Each row shows a feature’s SHAP value distribution as a violin plot (y-axis) against SHAP values (x-axis). Individual data points, consisting of 2,000 randomly sampled samples, are represented as colored dots, where the color intensity reflects the percentile rank of the feature value in the dataset, demonstrating the magnitude of the feature’s impact and the relative values of the features. A ranking of the top 20 features of the model. **b)** A ranking of the top 5 wearable features. **c)** A ranking of the top 5 lab features. **d)** A ranking of the top 5 ICU features. **e)** The aggregated feature importance by features in each group, scaled by the highest importance group. **f)** Dimensionality reduction (t-SNE)^29^ plotted for each averaged feature value group shown in e). **Statistical abbreviations**: Ct.: Count, Ent.: Entropy, Max: Maximum, Mean: Mean/Average, Min: Minimum, Sin.: Sine (trigonometrics), Skew: Skewness, Std.: Standard Deviation, Freq.: Frequency. **Technical abbreviations**: Embed: Embedding, PPG: Photoplethysmography, HRV: Heart Rate Variability, NN: Normal-to-Normal heart rate intervals, Act.: Activity: Standard Corsano feature generation, GT: GreenTeg Core wearable patch sensor, Q: Quality, RR: Respiration rate. **Clinical abbreviations:** CAM: Confusion Assessment Method, CRP: C-Reactive Protein (lab), Drn.: Drain, FiO2: Fraction of Inspired Oxygen, GGT: Gamma-Glutamyl Transferase (lab), HR: Heart Rate, ICU: Intensive Care Unit, LPS: Lipase (lab), MCH: Mean Corpuscular Hemoglobin (lab), OP: Intra Operative, SaO2: Arterial Oxygen Saturation (lab), SO2: Oxygen Saturation (lab), Temp: Temperature, WBC: White Blood Cell Count (lab), COHb: Carboxyhemoglobin (lab), K: Potassium (lab), MetHb: Methemoglobin (lab), Na: Sodium (lab).

### Evaluating ML-driven alarming strategy

To translate CASHEWS+ predictions into actionable clinical alerts, we developed an alarm strategy addressing the critical challenge of alarm fatigue, for example, in the well-monitored ICU where conventional threshold-based systems generate 100-1,000 alarms per patient per day, with up to 99.5% being non-actionable^32^. We prioritized three desirable properties: (1) low alarm burden operational within nursing capacity^33^, (2) similar precision at the same level of recall to maintain clinician trust, and (3) sufficient lead time for intervention while not alarming before any complication could have developed^34^. The source for the alarm system was the probabilities output by the model. As the basis of the system, we considered an 8h shift. We employed deactivating the alarm after a false positive alarm (*silencing*) for 8 hours^13^. The crossing of a threshold of alarms required in a sliding time window, known as *persistence*, was set to 3 times in the last 6 hours. The exact hyperparameters and methodology are described in the Methods section. The performance of the different alarm strategies was evaluated using a 24-hour prediction horizon, as this was seen as an appropriate lead time. We employed *precision*, *recall, and the number of alarms per patient per day* as evaluation metrics. Figure 5 demonstrates that the predicted probability for patients in the complication group generally increased to higher risk probabilities later in the progression of the stay. Moreover, we compare a sample patient with the REWS-based alarm and demonstrate that the alarms per patient per day (alarm burden), reduced significantly, while maintaining a similar precision at each level of recall, e.g., if an alarm burden of 2 is desired because of operational capacity, we obtain a recall of 0.9 and a precision of 0.20.

**Figure 5.**
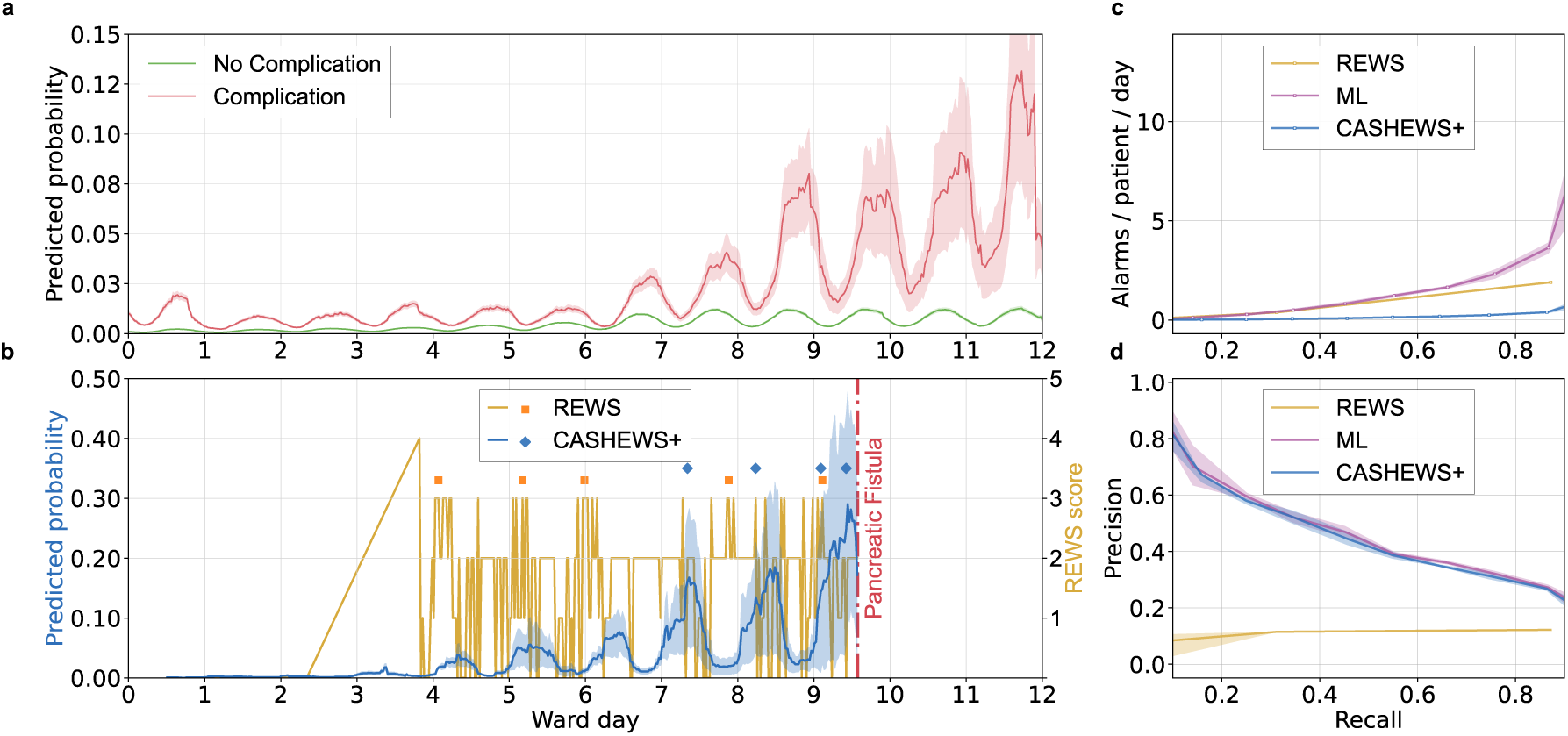
The implementation of CASHEWS+, a multi-modal wearable-enabled early warning system for surgical wards. **a)** ML predictions stratified by the complication and non-complication groups. **b)** An example patient stay from the first ward day to the end of their stay in the ward. REWS: Remote Early Warning Scoring^31^. CASHEWS+: Clinical ASsist Heuristic Early Warning System + Wearables. Individual points indicate alarms for the respective strategy. We chose the highest precision in the same recall bin that still produced at least one alarm. **c)** Alarm burden (alarms per patient per day) for the raw ML predictions with static threshold, the alarm model with blackout periods, a dynamic threshold, and a REWS-based dynamic alarm system with the same alarm strategy. **d)** The precision-recall curve according to Gao et al, ^28^ for the aforementioned alarming strategies.

## Discussion

Postoperative complications have been termed a “hidden pandemic,”^35^ reflecting their prevalence, lasting clinical impact, and the substantial economic strain on hospitals and health care systems. The CASSANDRA study introduces the concept of an AI-supported ward to identify and predict postoperative deep surgical-site infections through continuous, real-time analysis of multimodal patient data. To our knowledge, this is the first study to develop an AI-based monitoring system for general surgical wards, integrating data with high-resolution telemetric vital sign acquisition for predictive analytics. Our findings demonstrate the feasibility of digital patient representations to enable the early detection of postoperative complications, as evidenced by a model AUROC of 0.89. Moreover, prediction horizons of 6 and even 12 hours ahead of the current standard of care achieve a reasonable AUROC of 0.90 and 0.86, respectively. These results suggest that multimodal, real-time models could enable AI-supported wards, support the early detection of complications, and facilitate semi-autonomous clinical workflows.

Previous work on AI-based adverse event detection has primarily been confined to ICUs, where well-established continuous monitoring systems and the availability of large datasets facilitated retrospective analysis^13,36,37^. Most of these studies have targeted advanced events, such as circulatory failure^13^ or sepsis^14^, which typically represent only the terminal stage of a complication cascade, affecting fewer than 2% of patients. In contrast, the majority of postoperative complications occur on RWs, with early infectious events such as SSI-III, bile leakages, and pancreatic fistulas affecting up to 20% of patients and prolonging hospitalization by approximately 16 days^38^. Redirecting predictive monitoring to RWs, therefore, captures more frequent and actionable targets for enhancing perioperative safety. In this context, the implementation of telemedical sensors for continuous vital sign monitoring holds particular promise for both reducing nursing workload and enabling elaborate patient surveillance^39–42^. However, previous studies on monitoring strategies in this emerging field have predominantly evaluated predefined vital sign thresholds, simple rule-based heuristics, or existing early warning scores^43–45^. By contrast, we investigated a variety of sensor-derived signal analysis methods, including precomputed features from manufacturer-processed activity data, heart rate variability (HRV) and entropy features derived from raw PPG signals, and deep embeddings from PPG waveforms. The optimal wearable configuration improved model performance by 50% in terms of AUPRC, indicating that high-resolution sensor data is one of the most important modalities. The optimal setup was achieved with hybrid signal processing, which combined precomputed features with raw-signal embeddings, suggesting that high-dimensional signal structures reflect complex physiological interactions that extend beyond the scope of single-parameter monitoring. This finding is consistent with prior work demonstrating that PPG dynamics facilitate early sepsis detection, likely through the capture of microcirculatory alterations^46–48^. Moreover, HRV, temperature entropy, and respiration entropy were identified as the most substantial single feature contributors. In accordance, HRV has previously been linked to disease severity and prognosis in sepsis-associated states, as it reflects autonomic stress and neurocardiovascular dysregulation — both processes that typically precede clinical deterioration. Consistently, entropy measures reflect the “loss of physiological complexity” as an early marker of impending decompensation^49^. These findings underscore that dynamic complexity encodes early signs of physiological deterioration, making them usable for high-dimensional models. Embeddings derived from clinical documentation, including physicians’ and nurses’ free-text notes on examinations and bedside observations, contributed only marginally to model performance for the outcome of interest.

The model architecture is a crucial choice for any AI-driven system, as it impacts numerous characteristics of the resulting system. We trained several ML model architectures using the engineered segments to ensure optimal performance in our predictive setup: Deep Learning (DL) (Transformer^23^, GRU^24^, TCN^25^), Gradient Boosted Trees (GBT) (XGBoost^20^, LightGBM^21^, CatBoost^22^), Random Forest, and Logistic Regression. GBTs are theoretically at a disadvantage as they do not inherently capture temporal information, like DL architectures such as Transformers or Recurrent Neural Networks (RNNs). While DL is a popular technique for handling massive amounts of unstructured sequential and time-series data, we found that this approach did not outperform GBT models in our experiments. This is likely due to our robust, clinician-guided feature engineering, which performs historical aggregation and statistical extraction. GBTs are an effective feature selector^50^ that complements our multi-model, feature-rich data extraction approach. Moreover, recent ML literature^51–53^ has cast doubt on the superiority of DL and Large Language Models (LLMs) in many clinical prediction scenarios. Furthermore, using GBTs as prediction models has a key advantage: our setup runs on commodity hardware generally available in protected hospital environments, whereas DL is significantly more resource-and energy-intensive, as it requires the use of Graphics Processing Units (GPUs). Finally, model explanations are easier to extract than with deep learning, which is often a key evaluation criterion for certifying clinical models^54^.

Transforming predictive models into actionable alarm systems requires careful balancing of clinical utility with practical usability, which translates into a reasonable trade-off between sensitivity and the number of false alerts. Alarm fatigue is a well-known phenomenon in the ICU^55^. Our alarm strategies are based on predefined probability thresholds, temporal trends, and the frequency with which these criteria were met within a defined time window. These findings are consistent with everyday clinical practice, as our chosen endpoint is characterized by a slow progression and subtle symptoms that may develop over an extended period. Nevertheless, patients profit from early detection and countermeasures. However, since our model can trigger alarms up to 48 hours in advance, further investigation is needed to determine the extent to which these early alarms can be verified through diagnostic measures^56^. Complications detected this early may not yet be evident in, for example, a Computer Tomography (CT) scan. Our results further demonstrated that our ML models, combined with an optimized alarm strategy, substantially outperformed REWS. These findings suggest that structured, AI-generated alarms can detect subtle yet clinically significant deterioration early, while maintaining alarm frequency at a manageable level for clinical staff. An appropriate recall bin, number needed to evaluate, and prediction window need to be chosen by balancing the risk of excessive false alarms against the need to avoid missing complications, while also considering the available resources on a general ward.

This study has several limitations that may affect the generalizability of our findings. Despite being the largest reported cohort in surgical research using continuous vital sign monitoring, larger cohorts could enable more adaptable models. Moreover, as a single-center study conducted within our institution’s Enhanced Recovery After Surgery (ERAS) program^57^, the results reflect the implementation of strict, standardized post-operative procedures and an institutional culture with exceptionally low failure-to-rescue rates. While these factors contribute to optimal patient outcomes, they may limit the applicability of our findings to centers with different post-operative care standards or complication management approaches. Additionally, the standardized nature of our protocols, though beneficial for internal validity, may not reflect the variability in clinical practice encountered across different healthcare settings.

In conclusion, the concept of an AI-supported, semi-autonomous surgical ward directly addresses persisting and even aggravating challenges in current postoperative care. Today, many general wards rely on intermittent bedside assessments, often performed only a few times per day, which makes the early recognition of subtle complications highly unreliable. This problem is exacerbated by chronic staffing shortages, increasing patient volumes, and the growing complexity of surgical populations, which together limit the capacity of nurses and physicians to provide continuous surveillance^58^. As a result, many complications are detected late, at a stage when interventions are less effective and associated with higher morbidity, costs, and mortality. A semi-autonomous ward equipped with continuous telemonitoring and AI-assisted analytics can provide uninterrupted surveillance, prioritize patients at highest risk, and issue manageable alerts that reduce alarm fatigue. Such systems not only enhance patient safety through earlier interventions but also alleviate workload, standardize surveillance independent of staffing variations, and optimize resource use. In doing so, they offer a scalable solution to bridge the gap between the demand for surgical care and the limited availability of human resources, marking a necessary evolution in ward organization in modern healthcare. Despite the promising technical performance of multimodal continuous models, their utility can only be conclusively established through the evaluation of patient-centered outcomes: length of stay, comprehensive complication index, readmission rates, and mortality statistics. Future research is key to providing objective evidence of the system’s meaningful improvement in healthcare delivery and justifying its implementation in clinical practice.

## Reproducibility Statement

The GitHub repository, which contains an extensive adjustable preprocessing pipeline for our multimodal framework, can be found at: https://github.com/rvandewater/CASHEWS. The repository for performing reproducible experiments over multi-modal EHR data can be found at: https://github.com/rvandewater/YAIB. Due to the ethics statement signed during the study period, we can only provide summary statistics over the dataset and cannot fully release a version to the general public. Please see the section *’Ethics Approval and Patient Consent’* for more information.

## Data Availability

Due to the ethics statement signed during the study period, we can only provide summary statistics produced for the manuscript over the collected dataset. These data are available upon reasonable request to the authors. We cannot release a full version of all data to the general public. The GitHub repository, which contains an extensive adjustable preprocessing pipeline for our multimodal framework, can be found at: https://github.com/rvandewater/CASHEWS. The repository for performing reproducible experiments over multi-modal EHR data can be found at: https://github.com/rvandewater/YAIB.

## Acknowledgements and Funding

This work and some of the authors (R.P. van de Water, B. Pfitzner) were funded by the “Gemeinsamer Bundesausschuss (G-BA) Innovationsausschuss” in the framework of “CASSANDRA - Clinical ASSist AND aleRt Algorithms” (project number 01VSF20015). RPvdW received funding from the European Commission under the Horizon 2020 project INTERVENE (Grant agreement ID: 101016775). We acknowledge the work of Wenzel Schöning, Thomas Malinka, Robert Siegel, and Christian Denecke and their contribution to the clinical study. The authors acknowledge the Scientific Computing of the IT Division at the *Charité - Universitätsmedizin Berlin* for providing computational resources that have contributed to the research results reported in this paper. URL: https://www.charite.de/en/research/research_support_services/research_infrastructure/science_it/#c30646061

## Contributions

Author Abbreviations:

- R.P.v.d.W. -- Robin P. van de Water
- A.W. -- Axel Winter
- D.Z.L -- Daniela Zuluaga Lotero
- B.P. -- Bjarne Pfitzner
- L.F. -- Lara Faraj
- B.A. -- Bert Arnrich
- P.R. -- Patrick Rockenschaub
- I.M.S. -- Igor M. Sauer
- M.M.M. -- Max M. Maurer

These authors contributed equally: Robin P. van de Water (lead machine learning and AI specialist), Axel Winter (main medical specialist).

The project was supervised by Max M. Maurer.

A.W. and M.M.M. conceptualized and conducted the prospective trial. R.P.v.d.W, A.W., and M.M.M. conceptualized this work. A.W. supervised the study and data collection process; R.P.v.d.W designed and supervised the data integration, modality fusion, preprocessing and aggregation; R.P.v.d.W created the experimentation framework and modality data integration.

R.P.v.d.W. conceptualized the evaluation approach and created Figures 1-5 and all figures in the methods section and appendix. B.P. analyzed the wearable data and created the preprocessing pipelines. D.Z.L. and R.P.v.d.W. created the alarm system, as visualized in Figure 5, and corresponding statistics, with supervision. L.F. and M.M.M. provided feedback on the figures. P.R. provided supervision for the ML approaches. FK, WS, TM, and CD contributed to the verification of patient labels. R.P.v.d.W, A.W., D.Z.L., P.R., and M.M.M. wrote the manuscript with the assistance and feedback of all the other co-authors.

## Methods

### Study population

The ethics committee of Charité University Hospital provided formal approval (EA4/198/21) on 20 October 2021. The study was registered in the German Clinical Trial Register (DRKS - Deutsches Register Klinischer Studien) under the study ID DRKS00034973 (https://drks.de/search/de/trial/DRKS00034973). This study was funded by the “Gemeinsamer Bundesausschuss (G-BA) Innovationsausschuss” in the framework of “CASSANDRA – Clinical ASSist AND aleRt Algorithms” (project number 01VSF20015). Participants provided written informed consent for the use of their data in this study.

A total of 1,285 patients were prospectively enrolled in the study after providing written informed consent before the surgery, either in the outpatient clinic or on the surgical ward. These patients were consecutively identified upon admission for a planned major surgery, which fell within the predefined list of eligible surgical procedures.

Exclusions were applied as follows:

- Major surgery not performed (n=69): Patients who were scheduled for a major operation but ultimately did not undergo the procedure.
- Withdrawal from study (n=18): Patients who withdrew from the study after enrollment, either declining to continue participation or refusing to wear the required monitoring devices.
- Outcome adjudication (n=4): Patients who were excluded during the outcome adjudication process due to ambiguity in defining the presence or timing of postoperative complications. Their clinical trajectory was marked by different complications, and the censoring time point was not precisely defined.
- Lacking signal quality or amount (n=58): after retrieving, cleaning and analyzing the signal quality, if there was no PPG data at all, patients were excluded. For example, patients did not wear the device properly, which prevented it from getting a reading.

After exclusions, the final cohort included 1,136 patients for model development and evaluation. Patients were classified into four mutually exclusive groups based on the presence, severity, and timing of postoperative complications identified during their hospital stay. Patients are censored after reaching their first major endpoint and after a clinical prediction timeline of 20 days. Additionally, patients are censored after experiencing a major complication different from our endpoint.

The *target complications* are the most common highly granular complications after major abdominal surgery of the upper and lower gastrointestinal tract, major hepatobiliary and pancreatic surgery, are deep organ space infections (SSI-III), bile leakages^17^, and pancreatic fistulas^18,19^. These endpoints were combined because they all represent intra-abdominal pathologies associated with fluid collections that required intervention. These endpoints were subsumed under SSI-III for clinical reasons, reflecting everyday clinical practice.

Patients were classified into four mutually exclusive groups based on the presence, severity, and timing of postoperative complications identified during their hospital stay. Complications were first identified prospectively and later confirmed through structured chart review using strict clinical definitions based on the Clavien-Dindo (CD) classification:

- Group 1 – *No Complications:* Patients with no documented postoperative complications of any kind throughout their hospital stay.
- Group 2 – *Only Minor Complications (CD I&II):* Patients who had only minor complications during their stay—those not requiring significant interventions (e.g., conservative treatment only)—and no major complications at any time.
- Group 3 – *Target Major Complication First (CD III & IV)*: Patients who experienced at least one major complication, with the first major event matching the predefined target complication (e.g., intra-abdominal infection, anastomotic leak). This group reflects early clinical deterioration directly related to the study outcome of interest.
- Group 4 – *Other Major Complication First (CD III & IV):* Patients who also experienced a major complication, but their first major event was a different complication type than the predefined target. This group helps identify alternate deterioration patterns and competing risk events.

In cases where patients experienced both minor and major complications, the classification was based on the first major complication, as it marks the most clinically relevant point of deterioration. This logic ensures stratification by both severity and temporal sequence, aligning with the intended evaluation of early warning performance.

### Outcome annotation

The primary endpoint of this study (target major complication) was the occurrence of clinically relevant intra-abdominal complications. In this context, *clinically relevant* referred to complications that required an intervention—surgical, radiologic, or endoscopic. This definition corresponds to complications graded higher than Clavien–Dindo II, the most widely used classification system for postoperative complications among clinicians. Other major complications were defined as complications graded higher than Clavien–Dindo II, excluding surgical site infections grade III (SSI-III), bile leakage, and postoperative pancreatic fistula (POPF). A typical example would be postoperative respiratory failure requiring readmission to the intensive care unit and noninvasive ventilation (Clavien–Dindo IVa). Minor complications were defined as all complications with a Clavien-Dindo Grade less than III (i.e., pneumonia requiring antibiotic therapy). Table 2 contains a detailed definition of the primary endpoint.

Postoperative complications were identified prospectively by a visceral surgeon and a general physician through daily review of clinical data. To ensure accuracy, a second visceral surgeon independently performed a manual chart review after the availability of final laboratory and microbiological results following the interventions (typically within 2–5 working days). This multi-step process was implemented to guarantee rigorous and unbiased outcome ascertainment. These outcome assessments were conducted by physicians not involved in the treatment of study patients, ensuring objectivity. Wearable devices used to monitor vital signs had no visible display, making the data inaccessible to both patients and treating physicians. This ensured that clinical decisions, such as ordering CT scans, were based solely on the treating physicians’ clinical judgement, entirely independent of the study’s monitoring data, preserving unbiased outcome identification. In a subset of cases, a CT or MRI scan was performed before any intervention was indicated. The most common triggers for imaging, in descending order, were: highly abnormal laboratory values, Suspicious Fluid in the drainage, physical condition of the patient, and incidental findings on imaging performed for other indications.

We simplified the patient care process to four stages: patient admission, surgery, ICU stay, and general ward stay (Figure 6). We used a diverse range of data sources for modelling patients in our study. Several digital documentation systems were used to collect data. Admission information was recorded using a RedCAP^59^ database clinical trial application. The main clinical information system was I.S.H.Med^60^, which is primarily used to collect stay information and laboratory results. The ICU vital signs were sourced from the COPRA system^61^, which was used to collect vital signs, scores, and other ICU data. The wearable data was collected using the Corsano Band 1 (CardioWatch 287-1B) and Band 2 (CardioWatch 287-2B), wrist-worn devices that collect various physiological signals, of which the most important is PPG. Moreover, we measured the core temperature with the GreenTEG CORE device. The device data is collected using a Bluetooth device gateway mesh that transmits the data to a central server via the DeviceHub software^62^. Many features were renamed and aggregated after data exploration had determined that they represent the same measurement. Values were translated from German to English and standardized to a common unit of measurement. The timestamps are harmonized from the source time zone to the UTC+0 (GMT) time zone. Moreover, all features are strictly filtered to their respective time periods within each segment to ensure that no data leakage occurs; all features extracted from any events that occur in the current segment occur either during the segment or before it.

**Figure 6:**
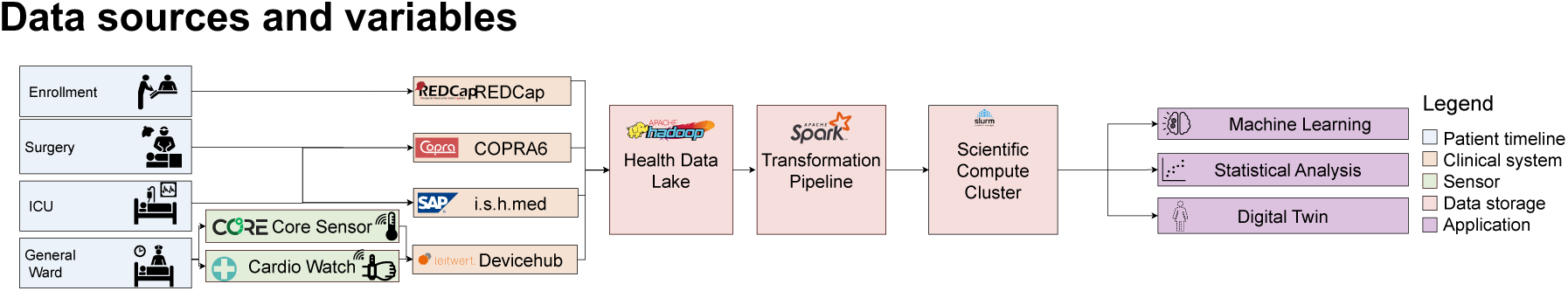
Complete data integration pipeline through each data management system.

**Figure 7:**
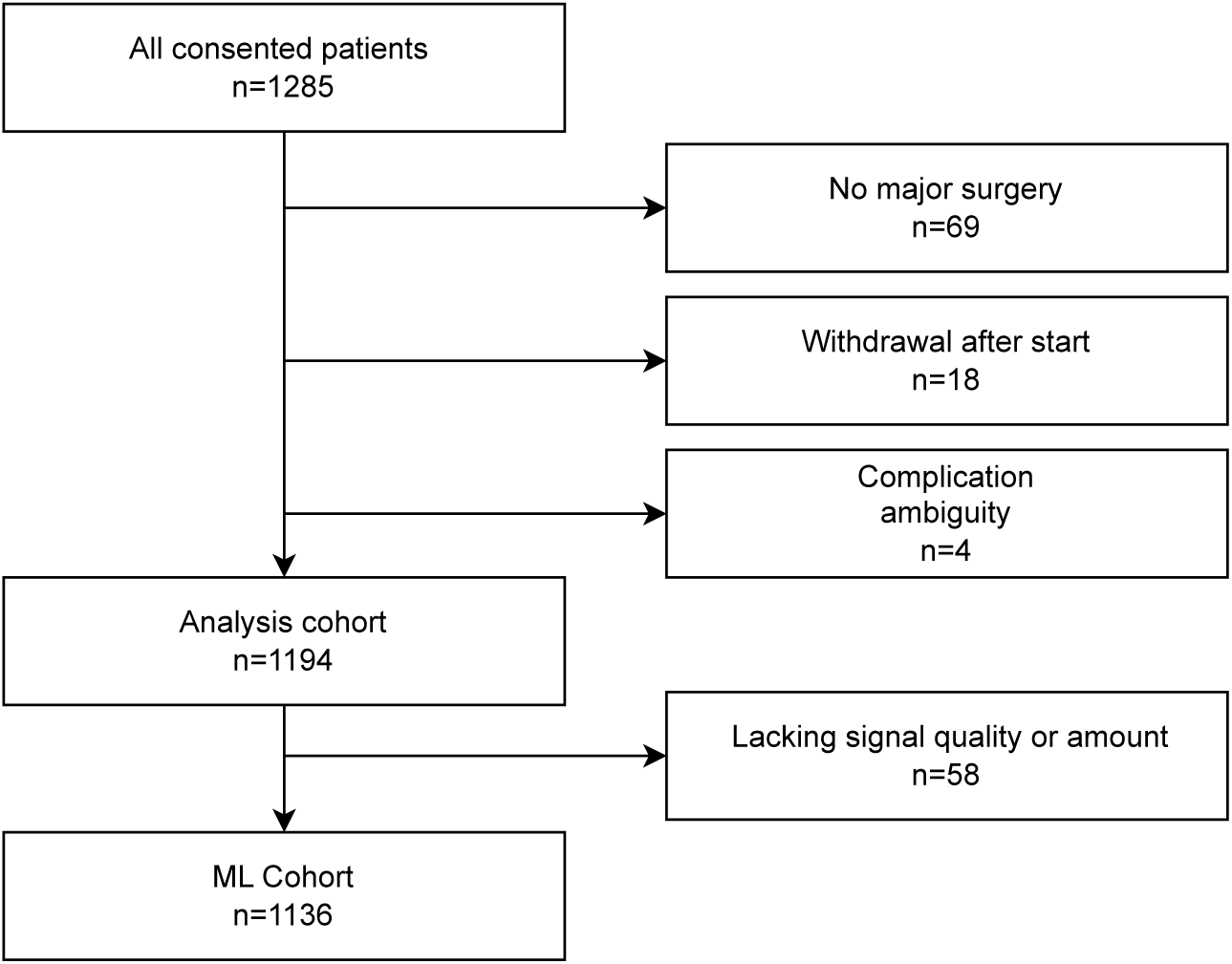
*Schematic exclusion criteria.* We use the analysis cohort for data exploration and analysis purposes and created a final ML Cohort that was used to train the CASHEWS+ models.

**Figure 7:**
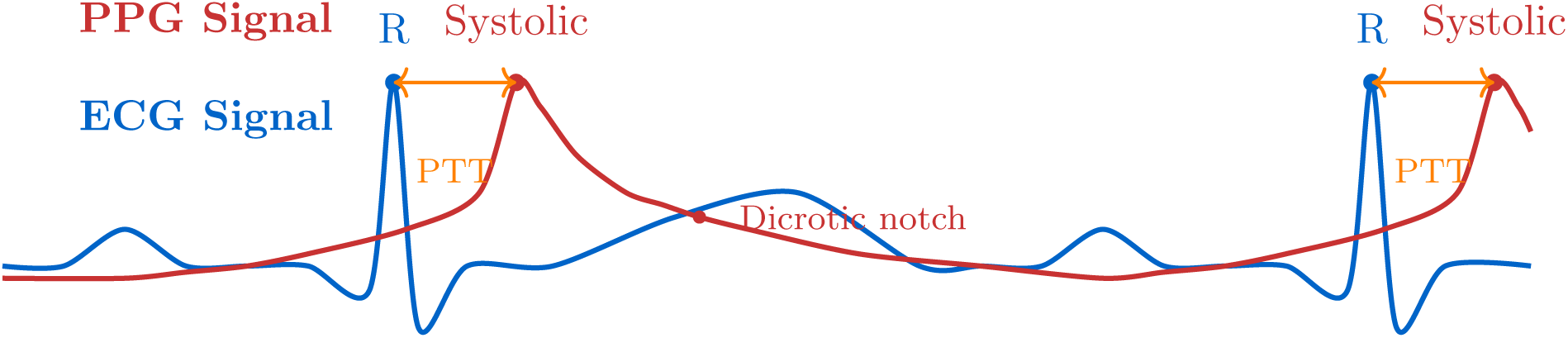
*Comparison of electrocardiogram (ECG) and photoplethysmogram (PPG) signals.* Upper trace (red): PPG signal showing sharp systolic peaks and dicrotic notches characteristic of arterial blood volume changes. Lower trace (blue): ECG signal showing sharp, biphasic QRS complexes representing electrical cardiac activity. Both signals are time-aligned but differ significantly in morphology, with the PPG reflecting hemodynamic changes and the ECG capturing electrical depolarization. The Pulse Transit Time (PTT) reflects the time it takes to detect the pulse at the PPG sensor, worn on the wrist.

**Figure 8:**
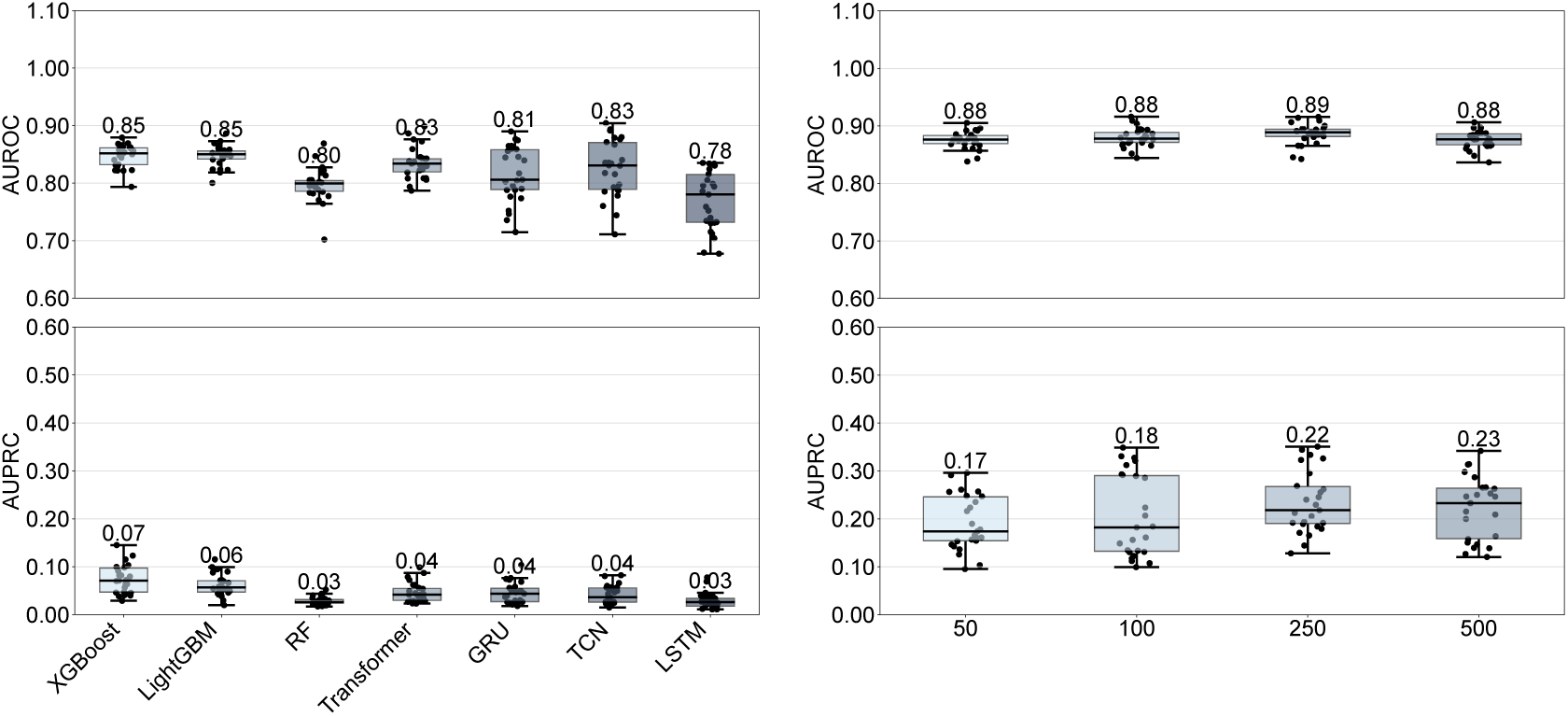
*Comparison of different model architectures. For* the non-gradient boosted models, a feature selection of the 500 most important was performed based on SHAPley values of an XGBoost experiment. Here, we did not perform the episode-level AUPRC as the trapezoid calculation distorted the results for results under 0.04.

### Domain-specific processing and fusion

We use early modality fusion^63^ to combine low-level features after performing initial preprocessing, feature extraction, and (where applicable) embedding. The segmented combined features were input to the modelling pipeline.

*Numerical ICU monitoring and lab values:* features were treated uniformly. They are aggregated per subject into 30-minute segments after experimentally determining the right segment length. For each segment, we extract the following statistical features: mean, minimum, maximum, standard deviation, count, skewness, entropy, median, and sum.

*Wearable data feature extraction:* The preprocessing pipeline first ensures that all PPG traces are standardized to 32 Hz and normalized per patient (by subtracting the mean and dividing by the standard deviation). Then, it splits the traces into contiguous stay-aligned segments and fixed-length windows (default: 60 s, 1920 samples) with optional overlap; windows with a standard deviation of zero are skipped. For embedding, MiniRocket applies deterministic, extremely sparse 9-length convolutions with only three nonzero taps (84 unique 3-tap combinations) at multiple log-spaced dilations to capture patterns across temporal scales suitable for 32 Hz sampling. For each kernel/dilation pair, a set of scalar bias thresholds is assigned using a low-discrepancy quantile sequence computed from empirical convolution outputs, resulting in diverse binary detectors. At transformation time, each dilated convolutional response is filtered with the proportion of positive values (PPV) operator (1 if the response exceeds the bias, otherwise 0) and then pooled by computing the temporal mean (proportion positive), yielding compact, shift-robust statistics per detector. Implementations include a Numba-accelerated CPU path that computes dilations, biases, and PPV pooling, and an optional output. Outputs are saved per segment as Parquet rows with feature columns named by index and segment length. This design emphasizes reproducibility, computational efficiency, and scale for large PPG cohorts sampled at 32 Hz.

The wearable data is processed in multiple ways to ensure that the high-resolution data can be used for the real-time alarm model. The raw PPG data is preprocessed with our pipeline:

1. Remove duplicates from the data and sort by datetime.
2. Segment data by identifying longer gaps (60s).
3. Resample signal to clean sampling rate (25/32Hz).

- Fill in gaps by cubic spline interpolation.

4. Apply 4th Order Chebyshev Type II Filter 0.5-5Hz
5. Detect motion artifacts (outside STD) from accelerometer data, set to NaN, and interpolate.

Further preprocessing is then applied to be able to feed the data into the segmentation pipeline efficiently:

1. Segmentation to 5-minute snippets
2. Calculation of HRV features using NeuroKit2^64^

a. Time Domain
b. Frequency Domain
c. Non-linear Domain
3. Reduction of 76 features down to 32 by aligning with features used also by e.g., Kubios^65,66^
4. Statistical aggregation in 1-hour and 6-hour (etc.) periods.

*Medications and clinical note features:* Clinical notes were recorded by nurses and physicians as part of routine care and are completely in German. We optimized the clinical texts by manually determining and replacing common abbreviations, anonymizing personal identifiers, and removing non-informative footers. Medications and fluid balance information from the ICU and general ward were manually harmonized by clinical assistance: measurements and values considered the same or very similar were treated as the same feature. The clinical notes and free-text medications are fed into a flexible, pre-trained, general multi-lingual embedding model (jina-embeddings-v3^27^) from the HuggingFace^67^ pre-trained model repository to obtain contextualized embeddings. The clinical notes dimensions were set at 64 dimensions, and the medications at 128 dimensions, after observing improved performance from these settings. These values were selected based on the model’s performance in preliminary experiments. The embeddings were then aggregated by taking the element-wise mean into the same segments as the numerical features. Table 4 provides an overview of the collected and recorded signals.

**Table 3:** Defining the endpoints precisely primary endpoint.

| Complication Type | Definition |
| --- | --- |
| **SSI-3** | a. Purulent drainage from an organ/space drain (e.g., closed suction drain, open drain, T-tube, or CT-guided drain).<br>b. Isolation of one or more organisms from fluid or tissue within the organ/space by culture or non-culture-based microbiological testing performed for clinical diagnostic or therapeutic purposes (excluding active surveillance testing).<br>c. Presence of an abscess or other evidence of organ/space infection detected by<br>– gross anatomic or clinical examination,<br>– histopathological assessment, or<br>– imaging studies demonstrating definite or possible infection. |
| **Bile leakage:** | Bilirubin concentration in the drain fluid at least 3 times the serum bilirubin concentration on or after postoperative day 3 or as the need for radiologic or operative intervention resulting from biliary collections or bile peritonitis. |
| **Postoperative Pancreatic Fistula (POPF)** | The patient is clinically unstable, often requiring management in the intensive care unit. The patient remains *nil per os* (no food or drink) with complete parenteral nutrition. CT imaging typically shows worrisome pancreatic fluid collections necessitating placement of a new drain. Antibiotic therapy and somatostatin administration are often required.<br>Since the primary endpoint required an intervention, only type C pancreatic fistulas were included in the primary outcome analysis. |

**Table 4:**
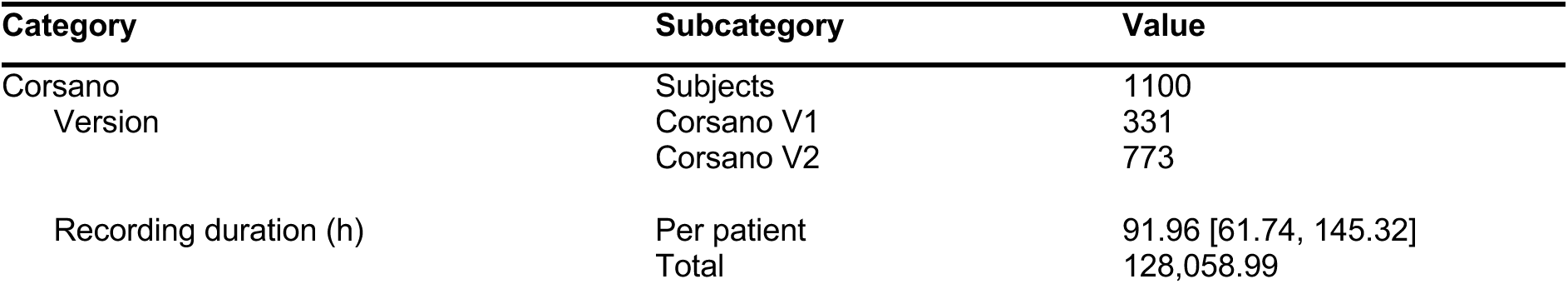
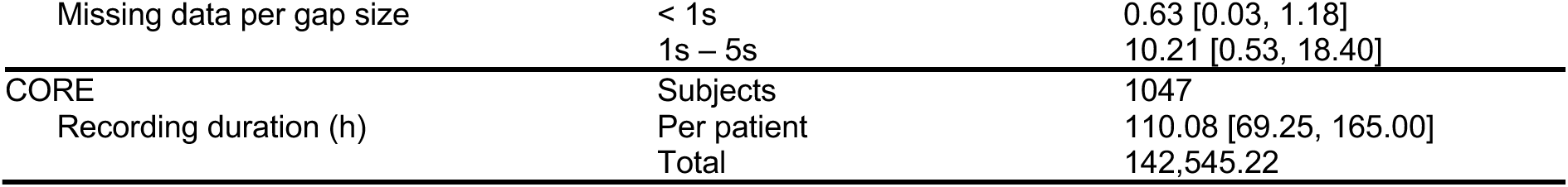
*Description of the collected wearable data.* When applicable, variables are summarized by median [IQR]. The gap sizes were chosen because of the nature of the signal; the Corsano outputs a high-resolution PPG signal, where small gaps can result in signal disturbances. The CORE device outputs the precomputed temperature.

*Data fusion and segmentation:* The data segmentation is a parallelized pipeline that segments the data into time intervals for monitoring across the patient’s trajectory (ICU and general ward). It aggregates information for a patient within an adjustable time period, while also truncating the data after the adverse event to prevent data leakage and produce a dataset suitable for training early warning systems (EWS). The feature extraction pipeline is designed to extract features from the segmented data. It encompasses a range of techniques, including statistical analysis, time-series decomposition, and domain-specific feature engineering, to capture the relevant information for each modality. Multi-horizon historical aggregation is performed to capture the temporal dynamics of the data, aggregating the data backwards at 30-minute, 1-hour, 6-hour, 12-hour, and 24-hour intervals.

We are able to flexibly: add new modalities; adjust segmentation periods and add/process new patient timelines, extract a range of adjustable feature sets based on statistical features and retrospective, summarized time-series data; modify prediction horizons and exclude patients without certain modalities; aggregate data from periods before the monitoring starts (pre-operative, operative, post-operative, ICU). Table 5 shows the resulting number of features per modality. To evaluate the temporal bin for our continuous analysis, data were aggregated into discrete time segments ranging from 7.5 minutes to 6 hours

**Table 5:** The number of features in each of the most granular modalities.

| Modality | N Features | Modality | N Features |
| --- | --- | --- | --- |
| Medications | 187 | Hour | 12 |
| Intra-OP Aggregated | 972 | Laboratory Values | 1998 |
| Comorbidities | 274 | PPG Embeddings | 2017 |
| Note | 98 | Wearable VS | 576 |
| ICU Fluids | 36 | Temperature | 108 |
| ICU Vital Signs | 372 | HRV Features | 195 |
| ICU Scores | 432 | **Total** | 7280 |

### Machine learning development and analysis

We expand an existing framework built for developing and benchmarking ICU models (see *Reproducibility and Adaptation to Different Clinical Settings*).

*Preprocessing:* In accordance with previous work, we performed runtime preprocessing steps to minimize data leakage between the training and test datasets.

The pipeline operates on three data segments: static patient characteristics, dynamic time-series measurements, and outcome labels. It processes training, validation, and test splits consistently using fitted transformations from the training data.

For static features, missing value indicators are generated across all static variables, followed by scaling and normalization procedures. Numeric variables undergo zero-fill imputation to address missing values, while categorical variables are processed through a two-step approach involving mode imputation to fill gaps, followed by label encoding to convert categories into numerical representations. The system creates missing value indicators for specified variables while respecting user-defined exclusions, then implements forward-fill imputation to propagate the last observed values forward in time. Any remaining missing values are addressed through zero-fill imputation, and the pipeline concludes by generating comprehensive historical aggregate features, including minimum, maximum, count, and mean values computed over time windows to capture temporal patterns in the data. The system supports recipe caching for transfer learning scenarios, allowing preprocessing pipelines to be saved and reloaded. All transformations are fitted on training data only and consistently applied to validation and test sets. This preprocessing approach addresses common challenges in data, including irregular sampling, missing values, and the need to combine static patient characteristics with time-varying physiological measurements while maintaining temporal relationships and preventing data leakage.

*ML architectures:* Several architectures were used: Deep learning (Transformers^23^, GRU^24^, TCN^25^), Gradient Boosted Trees (GBT: XGBoost^20^, LightGBM^21^, CatBoost^22^), Random Forest, and Logistic Regression. Adding more model architectures is easily possible due to the framework’s flexibility.

We selected the final classifier after thorough benchmarking, which aligns with earlier work indicating that boosted tree ensembles are a strong state-of-the-art classifier and often outperform Deep Learning models, such as Transformers^51,68^, especially when domain-guided feature extraction is employed.

This system produces probabilities for a temporal window of a patient’s stay. Each experiment was performed as a 5-times repeated 5-fold cross-validation to ensure the performance is balanced over the whole collected dataset. The test set was defined a 20 percent of subjects. The validation set was defined as 20% of the training subset. The patient timelines were split as a whole to ensure that data leakage between time windows could not occur (any patient window in the train set was not in the test set). Hyperparameter optimization was performed using predefined, publicly accessible configuration files. The traditional machine learning models were tested over 100 iterations of 5-fold cross-validation in a manner that the test set contained a unique subset of subject IDs each time. The Deep Learning models were instead trained with 50 iterations of tuning. Model calibration was performed but needs to be redone when training the final deployment model for the population.

### Explainability

The explainer plot visualizes the importance and impact of features in the machine learning model using SHAP (SHapley Additive exPlanations) values for the CASHEWS+ model with 250 features ranked by the highest SHAP contribution. The main visualization is a violin plot that shows the distribution of SHAP values for the top features across all predictions. Each feature is displayed with. The violin bodies represent the density distribution of SHAP values (wider sections indicate more predictions at that value). The individual data points, each comprising 1000 sampled values per violin, display the scaled magnitude of the feature value. In a SHAP plot, positive values indicate that the feature increased the prediction (higher risk), while negative values indicate that it decreased the prediction (lower risk). The features are ranked by their median absolute SHAP value, with the most influential features at the top.

To assess the contribution of different data modalities to model predictions, we aggregated SHAP values by their source modality. For each modality group, we computed the total absolute sum of SHAP values across all features and experiments to capture overall importance regardless of direction. To enable comparison across modalities with different feature numbers, we applied min-max scaling to transform aggregated values into a 0-100 range.

The plot includes a t-SNE (t-Distributed Stochastic Neighbor Embedding) visualization that projects the high-dimensional space of average feature values into 2D. This dimensionality reduction technique aims to demonstrate the distinctiveness of each feature.

### Alarm system

Alarm generation was based on a set of parameters optimized through hyperparameter tuning across 500 thresholds. The final configuration was selected with clinical feasibility in mind, striking a balance between early detection and practical implementation constraints. An 8-hour blackout period was applied after transfer to suppress early postoperative alarms. *Persistence* required three consecutive activations above the lower alarm threshold within a six-hour period to prevent transient fluctuations from triggering false alarms. This feature emphasized rising trajectories rather than isolated peaks, helping to identify sustained risk increases likely to precede complications. An 8-hour *silencing period*, where the alarm will not be triggered, follows each alarm to reduce alarm fatigue. To generate the precision–recall curve (Figure 5), the *lower threshold* was systematically varied to obtain recall bins while other parameters remained constant.

## Statistical methods

Unless specified otherwise, the solid lines and performance metrics presented in all figures and tables represent results from the test set of the’held out’ test data split described previously. The episode-based precision-recall curve was calculated according to an earlier work that explicitly described this evaluation method^28^. For box plots, the central line represents the median value, while the lower and upper boundaries correspond to the first quartile (Q1) and third quartile (Q3), respectively. The whiskers extend to show the first data point above Q1 – 1.5 × (Q3 – Q1) for the lower whisker, and the last data point below Q3 + 1.5 × (Q3 – Q1) for the upper whisker. Any points beyond these whiskers are marked as outliers. In violin plots, we used a bandwidth of 0.2 for the Gaussian kernel density estimation, ensuring the density distribution did not extend beyond the actual data range.

### Adaptation to different clinical settings

We adapted and thoroughly expanded an open-source ML benchmarking and development framework in active use by the Health AI community^37^ to work with any hospital stay timelines, not just ICU data. This provides a modular framework that allows researchers to define reproducible and comparable clinical ML experiments. Combined with a transparent preprocessing pipeline and extensible training code for multiple ML and deep learning models, YAIB enables unified model development, transfer, and evaluation. While working with our data collection framework, we added numerous features during the experimentation and development of our multimodal dataset, including modality selection, explainability (via Shapley) capabilities, and outcome horizon adjustment. These resources are again made available for the community to use in developing and benchmarking their own models, datasets, and preprocessing approaches at the open-source repositories of Yet Another ICU Benchmark (YAIB) (https://github.com/rvandewater/YAIB) and the Clinical ASsist Heuristic Early Warning System (CASHEWS) (https://github.com/rvandewater/CASHEWS).

### Analysis platform

The central Berlin Institute of Health’s central HPC cluster (Charité-HPC): with >5000 CPU cores, >80 GPUs, and >1.5PB high-performance data storage, was used. However, all our model training finishes within 48 hours except for hyperparameter tuning. Spark, Python with a.o. PyTorch, NumPy, Polars, Scikit-Learn, XGBoost 3.0.0, CatBoost 1.2.5, LightGBM 4.6.0, ImbLearn, and Optuna are part of the experiment platform.

## Appendix

**Table A:**
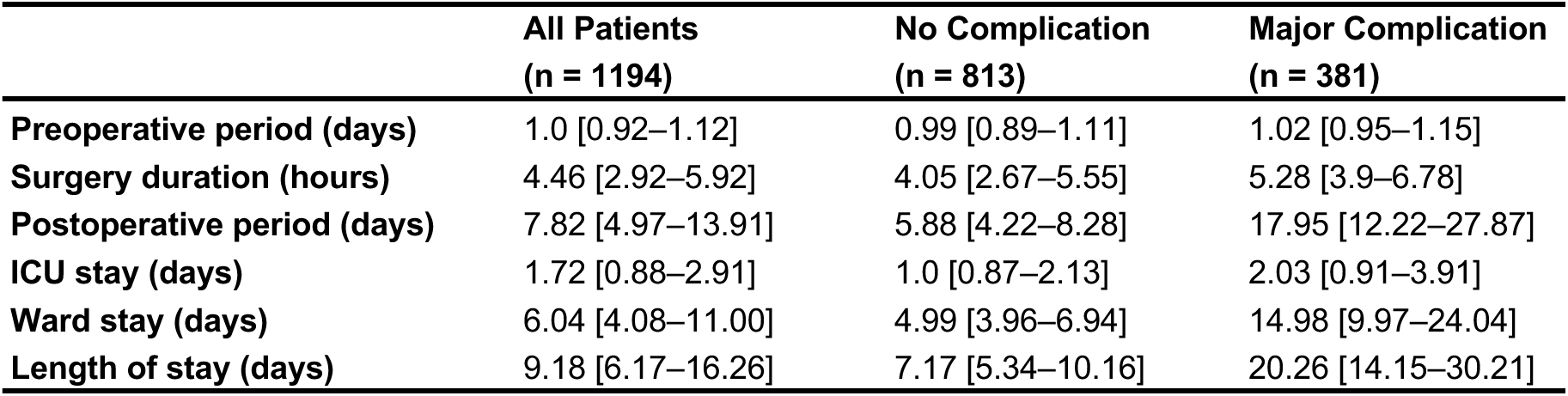
Intervals of all periods in the patient journey. All values shown as Median [Q1–Q3].

|  | All Patients<br>(n = 1194) | No Complication<br>(n = 813) | Major Complication<br>(n = 381) |
| --- | --- | --- | --- |
| Preoperative period (days) | 1.0 [0.92–1.12] | 0.99 [0.89–1.11] | 1.02 [0.95–1.15] |
| Surgery duration (hours) | 4.46 [2.92–5.92] | 4.05 [2.67–5.55] | 5.28 [3.9–6.78] |
| Postoperative period (days) | 7.82 [4.97–13.91] | 5.88 [4.22–8.28] | 17.95 [12.22–27.87] |
| ICU stay (days) | 1.72 [0.88–2.91] | 1.0 [0.87–2.13] | 2.03 [0.91–3.91] |
| Ward stay (days) | 6.04 [4.08–11.00] | 4.99 [3.96–6.94] | 14.98 [9.97–24.04] |
| Length of stay (days) | 9.18 [6.17–16.26] | 7.17 [5.34–10.16] | 20.26 [14.15–30.21] |

**Table B:** The modified REWS (Remote Early Warning Score) adapted from Van der Stam et al.^26^.

| Vital sign | Range | Score |
| --- | --- | --- |
| Heart rate (bpm) | < 40 or > 130 | 3 |
|  | 40–50 or 111–130 | 2 |
|  | 51–100 | 0 |
|  | 101–110 | 1 |
| Respiratory rate (breaths/min) | < 8 or > 30 | 3 |
|  | 9–11 or 21–30 | 2 |
|  | 12–20 | 0 |

**Table C:**
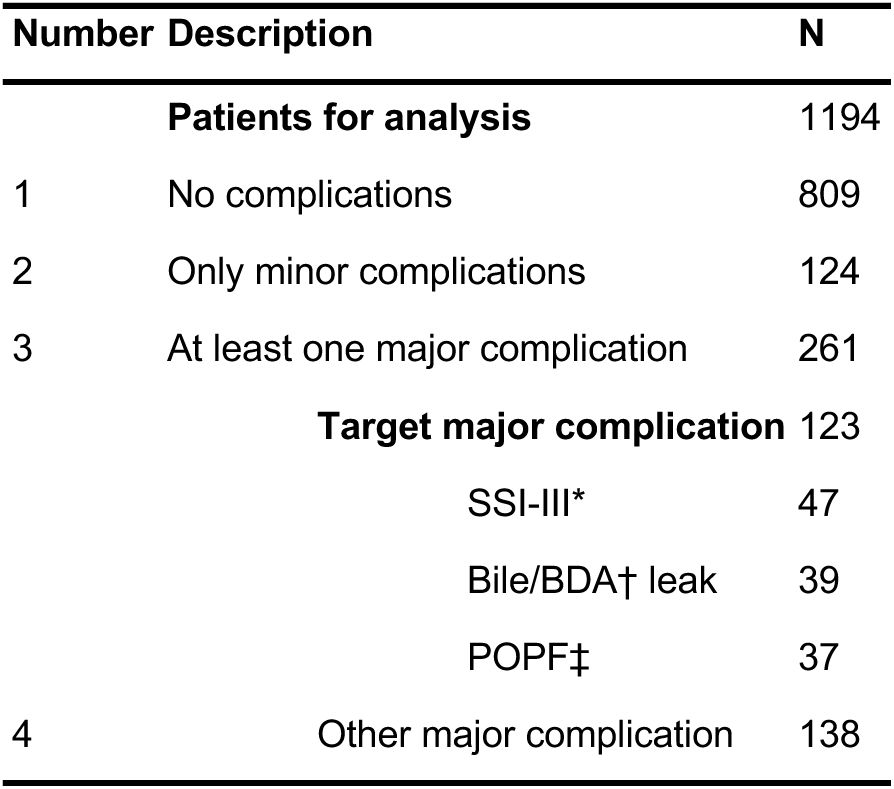
Description of the size of each group. Abbreviations: SSI-III = Deep surgical site infection, † BDA = Biliodigestive anastomosis, ‡ POPF = Postoperative pancreatic fistula.

| Number | Description | N |
| --- | --- | --- |
|  | **Patients for analysis** | 1194 |
| 1 | No complications | 809 |
| 2 | Only minor complications | 124 |
| 3 | At least one major complication | 261 |
|  | **Target major complication** | 123 |
|  | SSI-III* | 47 |
|  | Bile/BDA† leak | 39 |
|  | POPF‡ | 37 |
| 4 | Other major complication | 138 |

